# Mutation prevalence tables for hereditary cancer derived from multi-gene panel testing

**DOI:** 10.1101/19011981

**Authors:** Steven N. Hart, Eric C. Polley, Amal Yussuf, Siddhartha Yadav, David E. Goldgar, Chunling Hu, Holly LaDuca, Laura P. Smith, June Fujimoto, Shuwei Li, Fergus J. Couch, Jill S. Dolinsky

**Author notes:** Correspondence: Jill S. Dolinsky, MS, CGC, Hart, Steven N., Ph.D.

## Abstract

**Purpose:** Multi-gene panel testing for cancer predisposition mutations is becoming routine in clinical care. However, the gene content of panels offered by testing laboratories vary significantly, and data on mutation detection rates by gene and by panel is limited, causing confusion among clinicians on which test would be the most appropriate to order. Moreover, screening guidelines are not described in sufficient granularity to explain how differences in family, personal history, age, and other factors would affect the prevalence of finding a mutation in similar populations. The tool herein quantifies prevalence of mutations in hereditary cancer genes based on personalized clinical and demographic characteristics.

**Methods:** Using results from approximately 150,000 multi-gene panel tests conducted at Ambry Genetics, we built an interactive prevalence tool to explore how differences in ethnicity, age of onset, and personal and family history of different cancers affect the prevalence of pathogenic mutations in 31 cancer predisposition genes, across various clinically available hereditary cancer gene panels.

**Results:** Over 13,000 mutation carriers were identified in this high-risk population. Most of the cases were Non-Hispanic White (74%, n=109,537), but also provide an appreciable dataset for those identifying as Black (n=10,875), Ashkenazi Jewish (n=10,464), Hispanic (n=10,028), and Asian (n=7,090). The most prevalent cancer types were breast (50%), ovarian (6.6%), and colorectal (4.7%), which is expected based on genetic testing guidelines and clinician referral for testing.

**Conclusion:** The Hereditary Cancer Multi-Gene Panel Prevalence Tool presented here can be used to provide insight into the prevalence of mutations on a per-gene and per-multigene panel basis, while conditioning on multiple custom phenotypic variables to include race and cancer type. The tool can be found at https://www.ambrygen.com/prevalence-tool.

## Introduction

Between 5-10% of all cancers are associated with an inherited mutation in a cancer predisposition gene. The high rate of mutations has led to a plethora of academic researchers and genetic testing laboratories focused on defining the risk and prevalence for mutations in multiple genes and how they are associated with various cancers. In an attempt to provide some guidance into who should be tested for predisposition mutations, the National Comprehensive Cancer Network (NCCN) set out criteria to categorize individuals who are likely to contain a mutation in a predisposition gene - primarily based on an individual’s personal and family history of cancers. However, recent data has demonstrated limitations in these selection criteria for predicting who is more likely to have a positive pathogenic mutation^1-3^.

Historically, pre-test probability models have been the gold standard to assess the likelihood that an individual is a mutation carrier in *BRCA1*/2. These include BOADICEA^4,5^, BRCAPRO^6,7^, the Myriad II^8,9^, IBIS^10^, Penn II^11,12^, and Manchester^13,14^ models for breast cancers and MMRpro^15^ and PREMM^16^ for Lynch syndrome. These models, however, are limited to utility of predictions for *BRCA1* and *BRCA2*, as they are usually the only genes accounted for in these predictions due to the relatively low frequency of pathogenic mutations in other genes, however BOADACEA now also provides a pretest probability for *ATM, PALB2*, and *CHEK2* mutations^17^. These models were found to be reasonably accurate^18^, however, they were all derived from only a small number of cases or families which may present bias. For example, the Penn II model was derived from 169 women of whom 16% were positive for *BRCA1* mutations. Manchester, BRCAPRO, and BOADICEA were developed from 1121, 2713, and 2785 probands or families, respectively, of which ∼20% had pathogenic mutations in either *BRCA1* or *BRCA2*. The Myriad prevalence tables contain information initially derived from 10,000 consecutive cases through its clinical testing service; however the data has not been updated since 2010, and thus may no longer be representative of the population referred for hereditary cancer testing today.

While they have been incredibly useful, a key limitation to all pre-test probability models and existing prevalence tables/websites is the granularity at which they are published. For example, the Myriad tables only contain 2 populations, Ashkenazi and Non-Ashkenazi Jewish, with no information for mutation prevalence in Asian, Black or Hispanic populations. Moreover, family history information is limited to select combinations of breast and/or ovarian cancer personal and family history, even though there may also be histories of other cancers associated with hereditary cancer predisposition such as colorectal, endometrial, pancreatic, and prostate cancer. Some modeling tools can be overwhelmingly complicated or require downloading before running. If presented with insufficient numbers of exemplar data - or lack a strong statistical association for risk or outcome - then the model may not converge, failing to produce an accurate prediction. A platform providing a dynamic interface query based on substantial numbers of individuals tested in a more modern clinical setting based on multi-gene panel testing is likely to provide more precise estimations of mutation prevalence for a patient in a cohort of interest.

Simpler, interactive tools are making mutation prevalence data significantly easier to access. In 2018, Color Genomics released a website allowing quick perusal of genetic results from 50,000 individuals^19^. The user interface allows clinicians to estimate more refined mutation prevalence data using filtering criteria to better reflect the clinical characteristics of a given patient; however, the vast majority of tested individuals (n∼40,000) do not have a personal history of cancer, which may limit the utility of this tool.

Here, we describe the development and demonstrate the functionality of an open-access web-based tool which allows the end user to query mutation prevalence across 49 genes and 9 cancer indications with fine-grained control of demographic and clinical history factors. This tool represents data from 147,994 cases referred to Ambry Genetics for hereditary cancer testing, which is an order of magnitude larger than most of the datasets used for previous models. It also contains the largest database of underrepresented demographics referred for hereditary cancer genetic testing to include Asian, Black and Hispanic populations.

## Materials and Methods

### High-risk Patient Population

Study subjects included patients who underwent multigene panel testing through Ambry Genetics (Aliso Viejo, CA) between March 2012 and December 2016. Cases tested on the following panels were included: BRCAplus®, BreastNext®, CancerNext-Expanded®, CancerNext®, ColoNext®, GYNPlus®, OvaNext®, and PancNext®. Analysis of most genes on each panel consists of full gene sequencing of coding regions plus 5 base pairs into exon/intron boundaries (see **Table S1**.) with some exceptions^3^. Clinical histories were obtained from clinician-completed test requisition forms (TRFs), along with clinical documentation such as pedigrees and clinic notes, when provided. Prior research has demonstrated a high level of accuracy of such clinical information provided on TRFs compared to clinic notes and pedigrees, particularly for personal history of cancer^20^. Patients were asked to declare any family history of cancer, with specific categories for breast, ovarian, colorectal, pancreatic, thyroid, gastric, adrenal, prostate, and endometrial cancers. This study was deemed exempt from review by Western Institutional Review Board. Personal and family histories for breast, colorectal, melanoma, ovarian, pancreatic, prostate, thyroid, and uterine/endometrial were included if provided. Cases were grouped into one of five racial and ethnic categories based on self-report: and Non-Hispanic White, Black, Ashkenazi Jewish, Asian, or Hispanic. Only cases between 18-90 years old are included. For breast cancer, data from estrogen receptor (ER), progesterone receptor (PR), and human epidermal growth factor receptor 2 (HER2) statuses were included where available. Pathogenic mutations include variants with a classification of “pathogenic” or “likely pathogenic” based on a 5 tier variant classification scheme^21^.

### Interactive table application

Data were formatted into a custom R DataFrame (v. 3.3.3) object and loaded into an RShiny (v1.1.0) application. Filtering uses tidyverse (v.1.2.1), graphics with ggplot2 (v. 2.3.1). The application is located at https://www.ambrygen.com/prevalence-tool.

## Results

### Descriptive statistics of the cohort

Non-Hispanic Whites comprised the majority (74%) of the cohort. While breast cancer was the most prevalent cancer reported among cases (n=74,143, 50%), a large number of ovarian (n=9,768, 6.6%) and colorectal cancers (n=6,983, 4.7%) were represented as well. The median age of onset for any cancer was 56.2 years with a standard deviation of ± 13 years. Most of the patients tested were women, although 11,189 men were also tested, representing 7.4% of the total cohort.

### Family History

Some level of family history was reported for 143,448 cases. Not surprisingly, the most frequent reported family history of cancers was for breast (64%), followed by colorectal (28%), prostate (20%), and ovarian (18%). Mutation carriers were most prevalent in people with a family history of breast cancer (68%), followed by colorectal (29%), prostate (21%), and ovarian (19%). Interestingly, 5.9% of cases tested reported no family history of any cancer - 11% of which were found to carry a pathogenic mutation.

### Mutations

Of the 150,319 cases that underwent genetic testing, 13,401 carried pathogenic mutations-excluding heterozygous *MUTYH* carriers (14,475 mutations in total). Four genes were mutated in at least 1000 cases, including *CHEK2* (n=2,722), *BRCA2* (n=2,383) and *BRCA1* (n=2,282), and *ATM* (n=1,272). Another 18 genes (*PALB2, APC, MSH6, PMS2, BRIP1, TP53, MSH2, RAD50, MLH1, RAD51C, NBN, BARD1, CDKN2A, NF1,RAD51D, PTEN, MRE11A* and *CDH1*) had 100-800 pathogenic mutations. Pathogenic mutations in *MITF, MUTYH, FH, SMAD4, BMPR1A, FLCN, SDHB, SDHA, SDHD, STK11, EPCAM*, and *BAP1* were observed between 10 and 100 times. No pathogenic variants were identified in *CDK4 or TSC1*. Moderate risk mutations were included for *APC, BRCA1, CDKN2A, CHEK2, PMS2*, and *TP53*. In the case of *MUTYH*, only bi-allelic mutation carriers were considered. The c.952G>A (p.E318K) variant was the only tested variant for the *MITF* gene.

When comparing pathogenic mutation prevalence by gene, the overall number of mutations may be deceiving, as not all cases were tested for all genes, so we also describe mutations in the context of frequency and provide total number of cases tested on a by gene basis. Overall, *CHEK2* mutations were the most prevalent (2%), followed by *BRCA1, BRCA2, APC*, and *ATM* each being found in more than 1% of tested cases. By ethnic/racial population, the Ashkenazi Jewish cases presented with mutations in 18.1% of probands followed by 17.4% in Non-Hispanic Whites, 15.2% Hispanic, 14.5% African American, and 11.8% Asian.

### Exploration tool

Given the size, complexity, and value of this dataset, we built an interactive web-based tool to allow complex queries to better understand the landscape of inherited mutations in individuals at high risk for developing cancer (https://www.ambrygen.com/prevalence-tool). Users are able to select cases based on their age at first cancer diagnosis, personal history of different cancer types, family history of different cancer types, and ethnicities (Figure 1). The tool will return the prevalence of mutations for each gene, the prevalence of positive findings on nine different multi-gene panels offered through Ambry Genetics, as well as the distribution of both personal and family histories of patients after applying any of the above filtering criteria chosen by the user. As different genes are present on different panels, the number of mutations and the number of cases tested are also provided.

**Figure 1.**
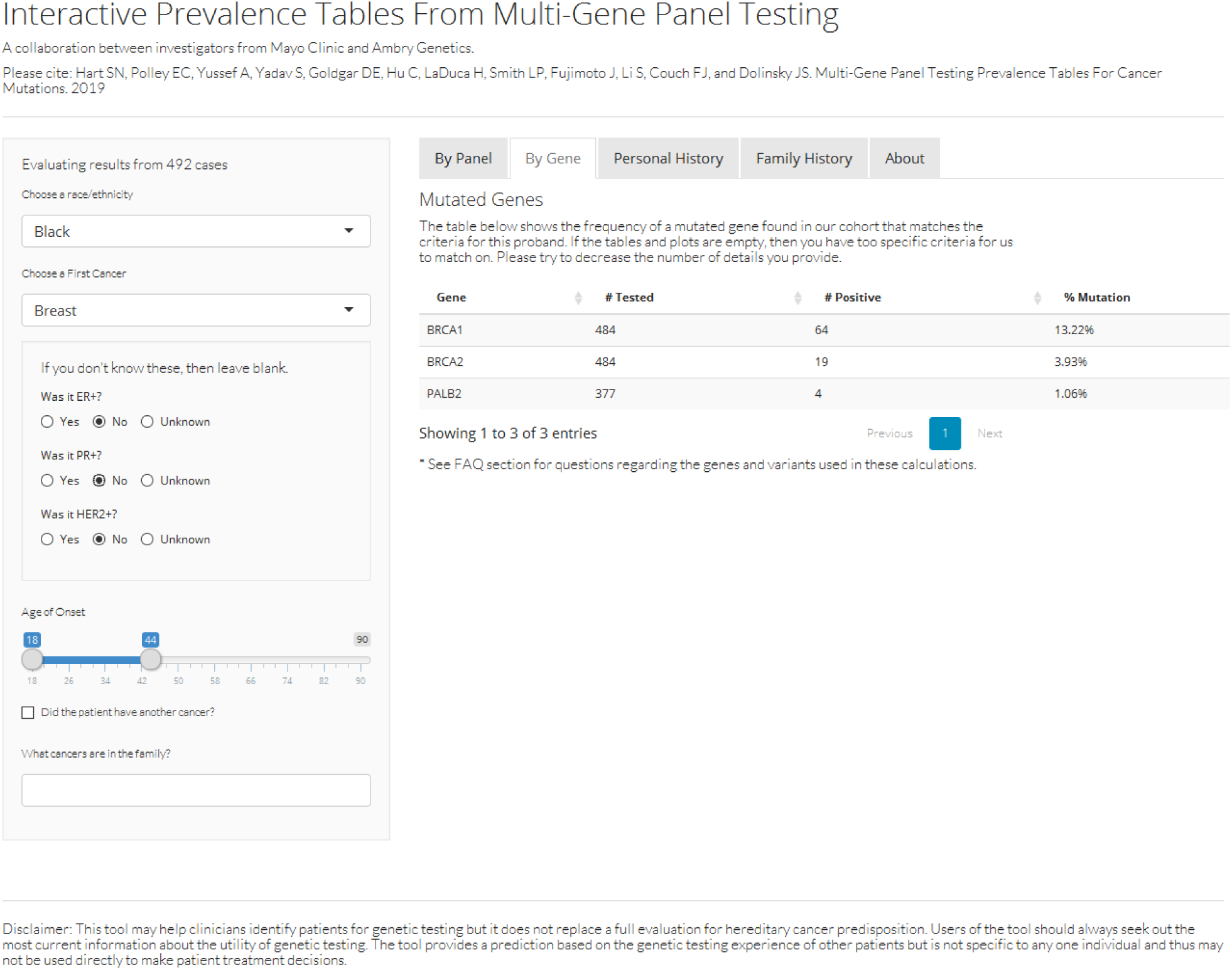
Screenshot of the interactive tool.

As a demonstration of the utility of the tool, we posed the following question: “How different are mutation frequencies in the *MLH1* gene from colorectal cancer cases with a family history of pancreatic cancer versus family history of prostate cancers?” To answer this question, the data were filtered for cases with a “First Cancer” of “Colorectal”, leaving the histology of those cases as “Unknown”, and then selecting the appropriate fields in the “Family History” section. Then, the number of positive mutations and the number of tested per gene are returned for all genes, including *MLH1*. The numbers of individuals tested and positive are returned for all genes, including *MLH1*, which in this case was 26/845 (3.08%) in pancreatic cancer family histories versus 22/1477 (1.76%) with a family history of prostate cancer. Feeding these values into a Fishers exact test confirm that pathogenic mutations were significantly higher in colorectal cases with a family history of pancreatic cancer (*p*=0.0149).

## Discussion

Understanding the prevalence of pathogenic mutations in patients at high risk for inherited cancers is of utmost clinical importance as it can dramatically differ based on ethnicity, family and personal history of cancer, age of onset, and genes tested. As such, results from the Hereditary Cancer Multi-Gene Panel Prevalence Tool could help inform test panel selection for clinicians and provide more personalized pretest anticipatory guidance for patients. For example, among breast cancer patients ages 40 to 70, mutation prevalence drastically increases from 4.6% with a 5-gene panel (BRCAplus) to 10.6% when selecting a 34-gene panel (CancerNext) and then again, albeit modestly, to 11.8% when using a 67-gene panel (CancerNext-Expanded). When choosing what panel is best for a patient, clinicians must weigh the relevance of the additional genes to the patient’s phenotype and varying incremental positive yield with the addition of a higher VUS rate as more genes are tested. They can further inform their decisions by referencing the gene-specific mutation prevalence rates within the tool. In addition, mutations in some genes have been seen fewer than 10 times, despite being sequenced in over 10,000 cases (*RET, GREM1, VHL, MEN1, MAX, TSC2, TMEM127*, and *SMARCA4*), suggesting a combination of limited involvement of these genes in the cancer histories seen in the majority of this cohort and rarity of mutations in these genes. For example, *SMARCA4* is associated with only a specific type of ovarian cancer, and *GREM1* mutations are limited to gross deletions/duplications and specifically associated with colorectal cancer.

While the Hereditary Cancer Multi-Gene Panel Prevalence Tool was primarily designed to support clinical decision making, it could also serve as a useful resource for researchers interested in studying a specific cohort. This tool would aid investigators in the study design process by allowing them to analyze broad trends and assess feasibility based on the size of a given cohort. This tool allows the flexibility to search the parameters of interest in an appropriate cohort rather than relying only on data breakdowns that others have previously published or asking targeted questions to the owners of the cohort data. For example, the tool shows that in cases under the age of 45, who had ER-positive breast cancer as their first cancer, mutations in the *CHEK2* gene are found in 4.3% of Non-Hispanic Whites compared to only 0.73% of Blacks. A researcher could assess whether the sample size by ethnicity is sufficient to address their research questions.

This tool does come with limitations. The data is based on a cohort of patients referred for hereditary cancer genetic testing due to clinical suspicion of hereditary cancer predisposition. Therefore, prevalence estimates may not be generalizable to the population at large, but rather should be viewed in the context of the clinical and family history provided within the tool. The clinical and demographic data is limited to that provided to the researchers and testing laboratory, although such a limitation is a reality in any cohort represented in a pretest probability model. In addition, while the size of the cohort contributing to this tool is orders of magnitude higher than that in most other currently available pretest probability models or tools, greater numbers of patients are still needed, particularly for ethnic minority populations, genes in which mutations are exceptionally rare, and queries for highly-specific patient characteristics.

Despite these limitations, this tool is representative of patients referred for hereditary cancer panels and is therefore highly relevant to current genetic testing practices. Continued efforts to update this tool and others like it will provide continuous benefits to patients and providers by supplying relevant information in a timely manner to support gene and panel test selection based on a patient’s personal and family history of cancer. Thanks to large scale data sharing from commercial and academic entities, it is now possible to explore complex queries that more accurately reflect the clinical experience through a simple web-based interface which draws upon data from large cohorts of patients recently referred for hereditary cancer multi-gene panel testing.

## Data Availability

The website is available at https://www.ambrygen.com/clinician/resources/prevalence-tool

## Acknowledgements

This work was funded by the Breast Cancer Research Foundation (BCRF #16-030), NIH Specialized Program of Research Excellence (SPORE) in Breast Cancer [CA116201], and the Mayo Clinic Center for Individualized Medicine.

---

**Table 1.** Summary age descriptions for each cancer type

|  |  | **Non-Hispanic**<br>**White**<br>**(N=109537)** | **Black**<br>**(N=10875)** | **Ashkenazi**<br>**Jewish**<br>**(N=10464)** | **Hispanic**<br>**(N=10028)** | **Asian**<br>**(N=7090)** | **Total**<br>**(N=147994)** |
| --- | --- | --- | --- | --- | --- | --- | --- |
| **Breast** |  |  |  |  |  |  |  |
|  | Unaffected | 51,341 | 4,042 | 5,538 | 4,816 | 2,868 | 68,605 |
|  | Mean Age of Onset (SD) | 50.3 (11.4) | 47.2 (11.2) | 52.6 (11.6) | 46.2 (10.7) | 45.6 (10.3) | 49.7 (11.5) |
|  | Range | 12.0 - 90.0 | 15.0 - 89.0 | 20.0 - 89.0 | 16.0 - 86.0 | 20.0 - 88.0 | 12.0 - 90.0 |
| **Ovarian** |  |  |  |  |  |  |  |
|  | Unaffected | 100,551 | 10,396 | 9,964 | 9,410 | 6,494 | 136,815 |
|  | Mean Age of Onset (SD) | 57.3 (13.4) | 54.5 (14.0) | 58.8 (13.7) | 51.8 (14.3) | 52.3 (13.6) | 56.7 (13.6) |
|  | Range | 5.0 - 90.0 | 14.0 - 86.0 | 11.0 - 88.0 | 16.0 - 86.0 | 17.0 - 88.0 | 5.0 - 90.0 |
| **Colorectal** |  |  |  |  |  |  |  |
|  | Unaffected | 103,169 | 10,268 | 10,112 | 9,467 | 6,729 | 139,745 |
|  | Mean Age of Onset (SD) | 50.0 (13.2) | 47.6 (12.0) | 52.6 (13.6) | 45.5 (12.6) | 45.2 (11.2) | 49.4 (13.1) |
|  | Range | 8.0 - 89.0 | 18.0 - 85.0 | 20.0 - 88.0 | 16.0 - 87.0 | 21.0 - 82.0 | 8.0 - 89.0 |
| **Uterine or Endometrial** |  |  |  |  |  |  |  |
|  | Unaffected | 105,734 | 10,651 | 10,167 | 9,725 | 6,892 | 143,169 |
|  | Mean Age of Onset (SD) | 54.3 (12.4) | 52.9 (13.2) | 57.4 (11.4) | 47.5 (13.1) | 48.9 (10.7) | 53.8 (12.5) |
|  | Range | 17.0 - 90.0 | 20.0 - 80.0 | 23.0 - 84.0 | 18.0 - 84.0 | 23.0 - 78.0 | 17.0 - 90.0 |
| **Pancreatic** |  |  |  |  |  |  |  |
|  | Unaffected | 108,215 | 10,773 | 10,257 | 9,945 | 7,026 | 146,216 |
|  | Mean Age of Onset (SD) | 60.8 (11.6) | 56.8 (11.9) | 64.7 (11.0) | 54.9 (12.9) | 53.7 (14.7) | 60.5 (12.0) |
|  | Range | 20.0 - 89.0 | 26.0 - 80.0 | 31.0 - 88.0 | 22.0 - 82.0 | 9.0 - 83.0 | 9.0 - 89.0 |
| **Thyroid** |  |  |  |  |  |  |  |
|  | Unaffected | 107,578 | 10,773 | 10,212 | 9,875 | 6,992 | 145,430 |
|  | Mean Age of Onset (SD) | 45.2 (13.8) | 46.6 (12.2) | 46.7 (13.8) | 45.8 (13.3) | 44.2 (11.4) | 45.4 (13.7) |
|  | Range | 8.0 - 89.0 | 21.0 - 78.0 | 6.0 - 75.0 | 16.0 - 81.0 | 14.0 - 74.0 | 6.0 - 89.0 |
| **Prostate** |  |  |  |  |  |  |  |
|  | Unaffected | 108,841 | 10,826 | 10,362 | 10,004 | 7,077 | 147,110 |
|  | Mean Age of Onset (SD) | 59.9 (8.6) | 58.9 (7.7) | 62.6 (7.9) | 61.0 (9.4) | 63.2 (8.9) | 60.2 (8.5) |
|  | Range | 34.0 - 85.0 | 39.0 - 78.0 | 45.0 - 81.0 | 46.0 - 84.0 | 50.0 - 82.0 | 34.0 - 85.0 |
| **Kidney** |  |  |  |  |  |  |  |
|  | Unaffected | 108,520 | 10,790 | 10,369 | 9,938 | 7,062 | 146,679 |
|  | Mean Age of Onset (SD) | 53.0 (14.8) | 51.4 (14.3) | 56.4 (12.2) | 47.9 (12.3) | 51.6 (11.0) | 52.7 (14.4) |
|  | Range | 1.0 - 87.0 | 6.0 - 77.0 | 27.0 - 79.0 | 2.0 - 74.0 | 31.0 - 74.0 | 1.0 - 87.0 |
| **Melanoma** |  |  |  |  |  |  |  |
|  | Unaffected | 106,848 | 10,863 | 10,191 | 10,000 | 7,080 | 144,982 |
|  | Mean Age of Onset (SD) | 47.7 (14.4) | 43.9 (16.2) | 49.3 (14.6) | 44.5 (14.9) | 43.4 (15.3) | 47.8 (14.5) |
|  | Range | 1.5 - 90.0 | 19.0 - 69.0 | 3.0 - 90.0 | 21.0 - 73.0 | 18.0 - 69.0 | 1.5 - 90.0 |

**Table S1.** Panels and genes used in this study.

| Panel | Genes |
| --- | --- |
| BRCAplus | BRCA1, BRCA2, CDH1, PALB2, PTEN, TP53 |
| BreastNext | ATM, BARD1, BRCA1, BRCA2, BRIP1, CDH1, CHEK2, MRE11A, MUTYH, NBN, NF1, PALB2, PTEN, RAD50, RAD51C, RAD51D, TP53 |
| CancerNextExpanded | APC, ATM, BAP1, BARD1, BRCA1, BRCA2, BRIP1, BMPR1A, CDH1, CDK4, CDKN2A, CHEK2, EPCAM, FH, FLCN, GREM1, MAX, MEN1, MET, MITF, MLH1, MRE11A, MSH2, MSH6, MUTYH, NBN, NF1, PALB2, PMS2, POLD1, POLE, PTEN, RAD50, RAD51C, RAD51D, RET, SDHA, SDHAF2, SDHB, SDHC, SDHD, SMAD4, SMARCA4, STK11, TMEM127, TP53, TSC1, TSC2, VHL |
| CancerNext | APC, ATM, BARD1, BRCA1, BRCA2, BRIP1, BMPR1A, CDH1, CDK4, CDKN2A, CHEK2, EPCAM, GREM1, MLH1, MRE11A, MSH2, MSH6, MUTYH, NBN, NF1, PALB2, PMS2, POLD1, POLE, PTEN, RAD50, RAD51C, RAD51D, SMAD4, SMARCA4, STK11, TP53 |
| ColoNext | APC, BMPR1A, CDH1, CHEK2, EPCAM, GREM1, MLH1, MSH2, MSH6, MUTYH, PMS2, POLD1, POLE, PTEN, SMAD4, STK11, TP53 |
| GYNPlus | BRCA1, BRCA2, BRIP1, EPCAM, MLH1, MSH2, MSH6, PALB2, PMS2, PTEN, RAD51C, RAD51D, TP53 |
| OvaNext | ATM, BARD1, BRCA1, BRCA2, BRIP1, CDH1, CHEK2, EPCAM, MLH1, MRE11A, MSH2, MSH6, MUTYH, NBN, NF1, PALB2, PMS2, PTEN, RAD50, RAD51C, RAD51D, SMARCA4, STK11, TP53 |
| PancNext | APC, ATM, BRCA1, BRCA2, CDKN2A, EPCAM, MLH1, MSH2, MSH6, PALB2, PMS2, STK11, TP53 |

